# The 14-Day Incidence and Risk Factors of Gastrointestinal Anastomotic Leak Among Adult Patients in Mulago Hospital

**DOI:** 10.1101/2024.11.24.24317847

**Authors:** A PROSPECTIVE COHORT STUDY, Isaac Omare, Ronald Mbiine, Brian Kasagga, Wilberforce Musoga Kabweru, Paul Okeny

**Affiliations:** Makerere University Faculty of Medicine: Makerere University College of Health Sciences; Department of Surgery: Mulago National Referral Hospital

**Keywords:** Anastomotic leak, Gastrointestinal Surgery, Incidence

## Abstract

**Introduction:** Anastomotic leak is a dreaded complication following gastrointestinal (GI) anastomotic surgery. It increases morbidity and mortality of patients undergoing GI surgery. However, there is paucity of data on the incidence and risk factors of anastomotic leak following gastrointestinal surgery in Uganda. The main objective of this study was to determine the incidence and risk factors of anastomotic leak following gastrointestinal surgery in Mulago Hospital.

**Methods:** We consecutively recruited adult patients admitted to the general surgery wards in Mulago Hospital, 24 hours following gastrointestinal surgery. The recruitment process started on 22^nd^ April 2024 and ended on 11^th^ July 2024. The participants provided informed written consent prior to enrolment into the study. Patient related factors including preoperative anemia, preoperative albumin level, and ASA status were recorded on entry, while the outcome (anastomotic leak) was obtained upon 14 day’s follow-up. Data were analysed using SPSS version 26. Multivariate logistic regression was used to determine the independent risk factors for anastomotic leak, p< 0.05 was considered statistically significant.

**Results:** Eighty-five participants were studied. The incidence of anastomotic leak was 8.2% (n=7). Hemoglobin level less than 10 g/dl was the only independent predictor for anastomotic leak in this study. (RR, 8.15; 95% C.I, 1.16 - 57.48; p=0.035).

**Conclusion:** The incidence of anastomotic leak in Mulago National Referral Hospital was low. Low Hemoglobin (Hb<10g/dl) was associated with increased rates of anastomotic leak.

## Introduction

Anastomotic leak is one of the most feared complications following GI anastomotic surgery (Girard et al., 2014). It is associated with increased in-hospital morbidity, and mortality. (Kassahun et al., 2022)

Anastomotic leak following colorectal surgery in Mulago National Referral Hospital was at 12% (Nathani, 2022). This statistic is slightly higher compared to the regional rates which are below 10%(Kassahun et al., 2022; Zemenfes & Tamirat, 2019). Studies globally have reported various risk factors for gastrointestinal anastomotic leak such as low Hb level, higher ASA score and low serum albumin levels.(Brisinda et al., 2022; Mori et al., 2023). However, there is paucity of data on the risk factors for anastomotic leak following gastrointestinal surgery in Uganda, with only one retrospective cross-sectional study looking at colorectal surgery in the literature (Nathani, 2022).

Identifying gastrointestinal surgery patients at high risk for anastomotic leakage will facilitate informed preoperative patient counselling and optimisation of patients preoperatively. This could potentially lead to improvement in the quality-of-care for patients as well as reduction in morbidity and mortality for these surgery cases in our setting.

## Materials and Methods

This was a prospective descriptive study conducted over a period of 3-months in Mulago Hospital, Kampala, Uganda. We consecutively recruited adult patients admitted to the general surgery wards in Mulago Hospital, 24 hours following gastrointestinal surgery. The recruitment process started on 22^nd^ April 2024 and ended on 11^th^ July 2024. The participants provided informed written consent prior to enrolment into the study. Patient related factors including preoperative anaemia, preoperative albumin level, and ASA status were recorded on entry, while the outcome (anastomotic leak) was obtained upon 14 day’s follow-up. The study population consisted of 85 patients aged 18 years and above admitted for elective and emergency surgery. Patients who had protective proximal stoma were excluded. The patients were assessed postoperatively for signs of anastomotic leak. 85 patients underwent Gastrointestinal anastomosis in the Hospital Theatres. The patients were assessed on day 3, day 7 and day 14 for clinical signs and symptoms of anastomotic leak. Theatre notes were reviewed to check for the indications for surgery, method of anastomosis used, and surgical technique used and Biochemistry laboratory results. Anastomotic leak was noted as a clinical diagnosis confirmed upon relaparotomy or imaging. The Follow-up of the patients was done up to 14th postoperative day. Patients were reviewed after 2 weeks in Surgical Outpatient Department. Telephone contacts were used, where possible. Patients who developed leaks were linked to the surgical team for appropriate management

Statistical data analysis was conducted using the SPSS version 26.0 software. A chi-square/Fisher’s exact test was used in testing association of categorical variables. The quality control was ensured by making the principal investigator carryout all the postoperative assessment, clinical examinations and measurements of parameters using standard international units (SI) to avoid inter-observer error. Laboratory investigations were done by the same method and in standard unit.

## Results

### Participants characteristics

Over the 3-month study period, 85 participants were recruited. The mean age was 44.8 (SD±16.4) with a range between 18 to 85 years. Majority (n= 49) of participants were males. Based on the indication for surgery, most patients (n=80) presented with diseased bowel. In addition, majority of patients had Hemoglobin concentration of ≥10g/dL, (n =70). More than half of the patients (n= 45) had an ASA score of ≥3.5. Most of the operations were elective, (n=43). Open surgeries (n=79) were the frequently most done in this study. Small bowel anastomoses were done in more than half of the patients, (n= 49). Less than a quarter of the patients (n=16) received blood transfusion intraoperatively. (Table 1).

**Table 1:**
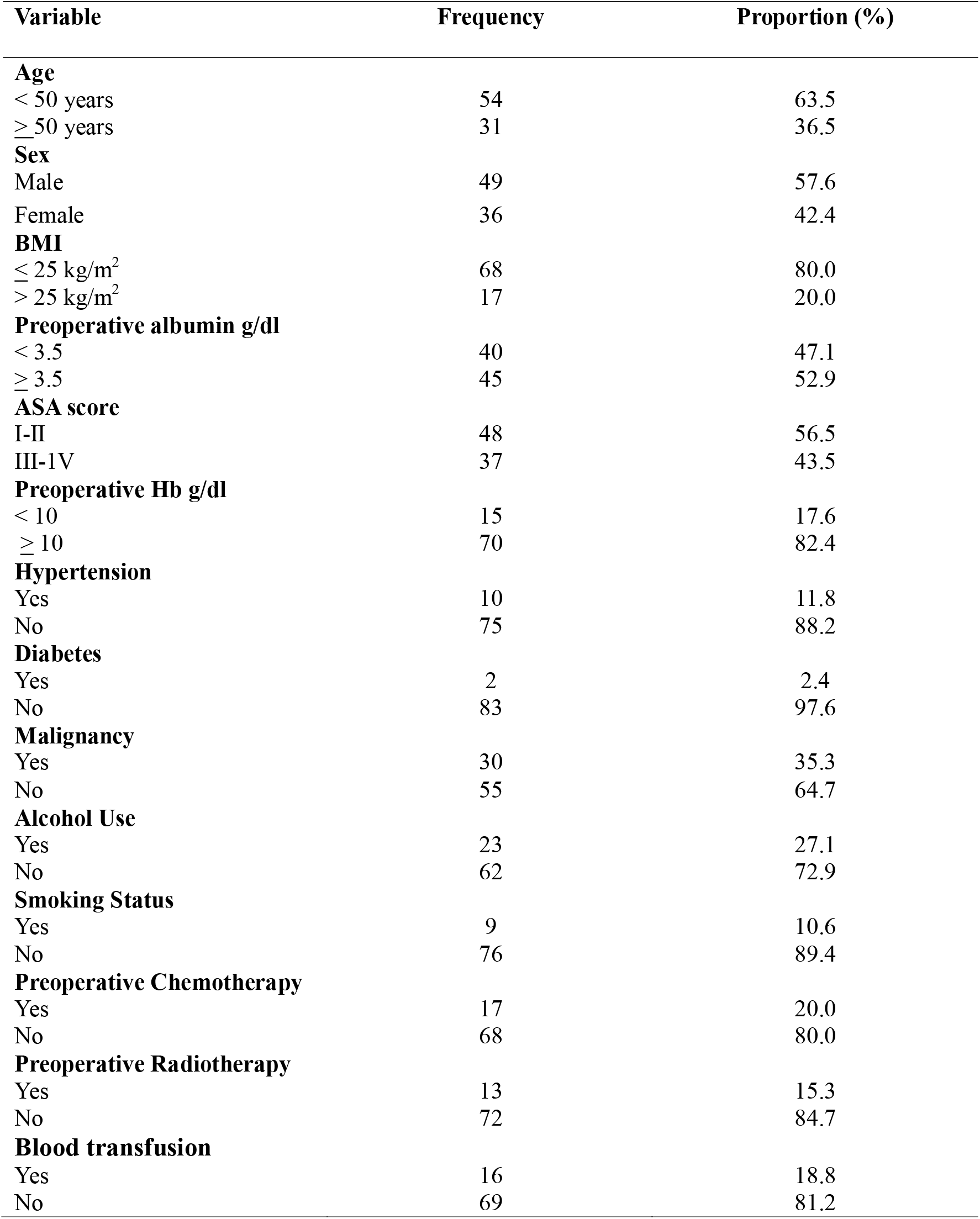
Characteristics of patients who underwent gastrointestinal anastomosis.

### Pattern of occurrence of anastomotic leak among patients that underwent gastrointestinal surgery at MNRH

In this study, the overall incidence of gastrointestinal anastomotic leak was 8.2% (n=7); (Fig. 2) Incidence by level of anastomosis was as follows; 8.3% in small bowel anastomoses as compared to 8.1% in large bowel anastomoses. There were (n=5) males and (n=2) females who developed anastomotic leak following gastrointestinal surgery. Overall, leaks were diagnosed at a mean of 5.3 days (range 3–8) postoperatively.

### Bivariate Logistic Regression Analysis

Following the variance inflation factor analysis for multicollinearity, type of surgery and with theatre used were collinear, therefore dropped, and Preoperative albumin g/dl, Preoperative Hb g/dl, and Cadre of Surgeon (Specialist vs SHO), were included in the multivariate regression model. Confounders were adjusted for following multivariate logistic regression analysis method. Out of the five significantly associated factors for anastomotic leak in bivariate analysis, only Preoperative Hemoglobin <10g/dl was shown to be an independent determinant of anastomotic leak after intestinal anastomosis. (RR, 8.15; 95% CI, 1.16 - 57.48, p = 0.035); (Table 3)

**Table 2:**
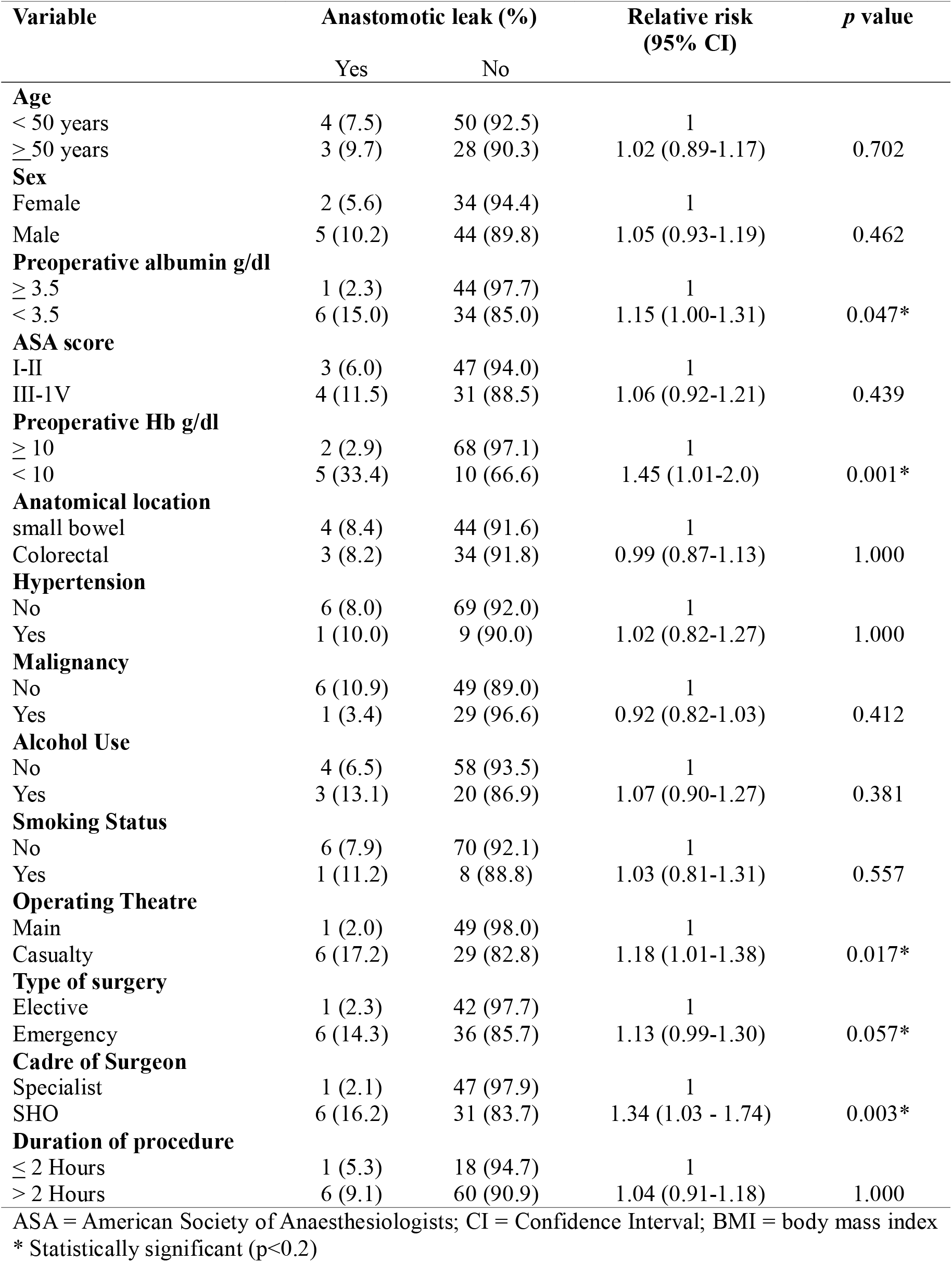
Bivariate analysis of patient and operative factors to determine association with intestinal anastomotic leaks.

**Table 3:** Multivariate logistic regression analysis of the independent risk factors of intestinal anastomotic leak.

| Variable | Frequency (%) | Relative Risk (RR) (95% CI) | p-value |
| --- | --- | --- | --- |
| **Preoperative Hb g/dl** |  |  |  |
| ≥ 10 | 70 (82.4) | 1 |  |
| < 10 | 15 (17.6) | 8.15 (1.16- 57.48) | 0.035* |
| **Preoperative albumin g/dl** |  |  |  |
| ≥ 3.5 | 45 (52.9) | 1 |  |
| < 3.5 | 40 (47.1) | 2.31 (0.19- 27.78) | 0.511 |
| **Cadre of surgeon** |  |  |  |
| Specialist | 48 (56.5) | 1 |  |
| SHO | 37 (43.5) | 1.95 (0.88- 4.34) | 0.100 |

Hb – Hemoglobin. \* Statistically significant (p<0.05)

**Figure 1:**
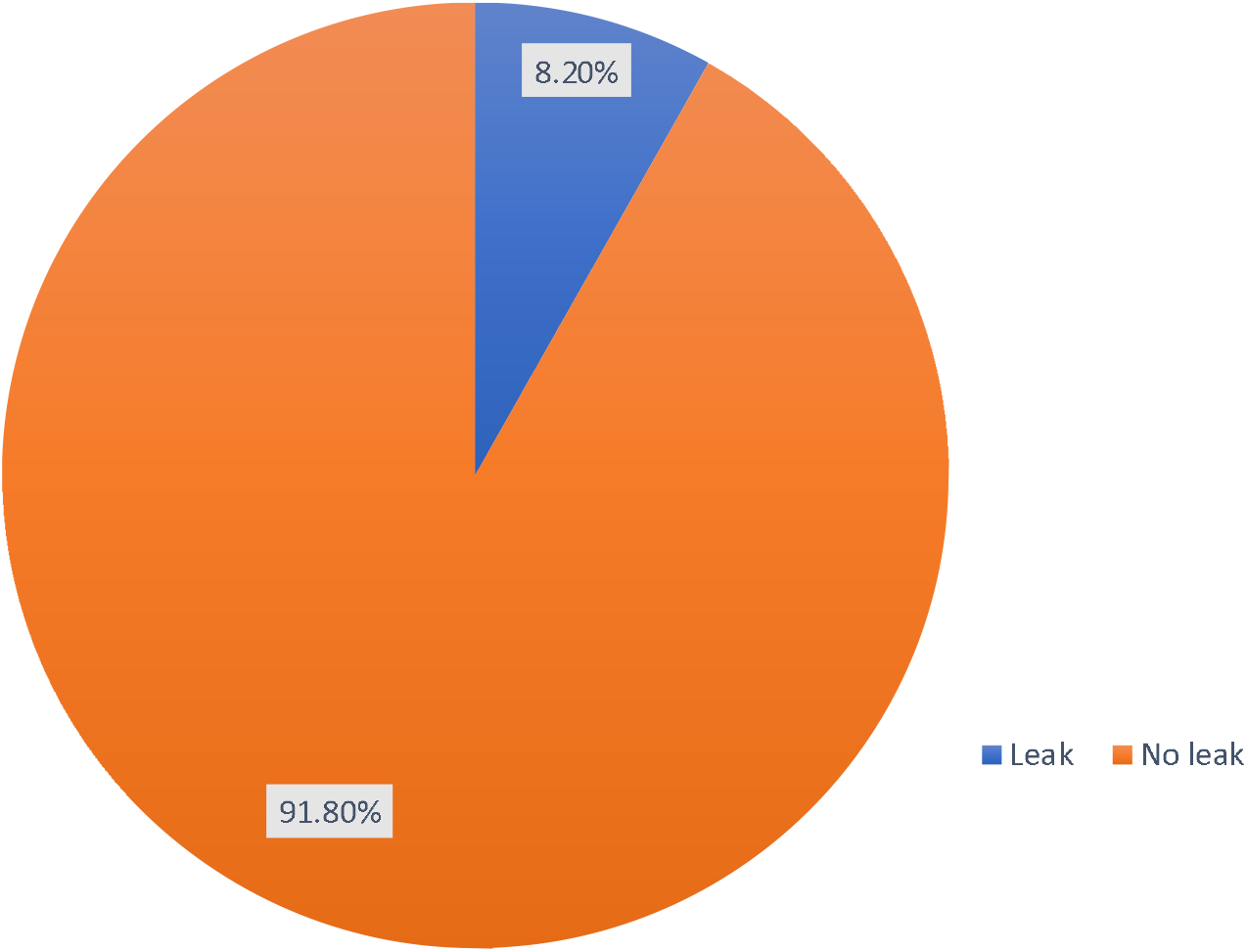
Anastomotic leak rate following GI anastomosis at Mulago National Referral Hospital.

## Discussion

### Basic characteristics of the study population

In this study, 57.6% of the participants recruited were male, (n=49) and 42.4% were female, (n=36). Males present more with GI surgical conditions requiring resection and anastomosis as compared to the female counterparts (Amone et al., 2020). The mean age of the participants was 44.8 (SD±16.4) with a range between 18 to 85 years. The wide range of age points out that gastrointestinal conditions occur in all age groups.

### Incidence of anastomotic leak

The incidence of anastomotic leak in our study was 8.2%. This is consistent with previous studies, which reported rates of 8.5% (Awad et al., 2021; Huisman et al., 2022). This is likely due to the similarity in study designs of these studies, whereby all of them were prospective in nature.

The incidence of leak in the current study is, however, higher than that reported by Wismayer (2021) who found the incidence of 7.7%. This is due to the differences in the study populations and study setting; his study was conducted in northern Uganda and he only studied patients with sigmoid volvulus whereas our study looked at both small and large bowel surgical conditions.

In contrast, several other studies have found higher incidences of anastomotic leak ranging from 9.2% to 16.8% (Banda, 2017; Kassahun et al., 2022; Zemenfes & Tamirat, 2019). This is likely due to differences in study designs in these studies compared to our study. Banda (2017) conducted a retrospective cohort study in Malawi whereas Kassahun et al. (2022) and Zemenfes and Tamirat (2019) conducted retrospective cross sectional studies in Ethiopia; compared to our study; a prospective cohort study, hence the above differences in anastomotic leak.

### Factors associated with anastomotic leak among study participants

Although age wasn’t a significant risk factor for anastomotic leak following gastrointestinal surgery, majority of leaks, 9.7%, happened in the older age group (≥50 years). The results of this study are contrary to those found by Kassahun et al. (2022) in Ethiopia; who reported age as a significant risk factor for anastomotic leak. This is due to the differences in the sample sizes. Kassahun et al. (2022) study; had a sample size of 411participants whereas our study recruited 85 participants.

Similarly, gender wasn’t a significant factor for anastomotic leak following gastrointestinal surgery. This is contrary to the study done by Sakr et al. (2017), which revealed that male gender was significantly associated with AL. This variation could be explained by the difference in the sample sizes of these studies. Sakr et al. (2017), recruited 224 participants whereas our study recruited 85 participants.

ASA class ≥3 was not a statistically significant risk factor for anastomotic leak in this study. This is contrary to the study done by Gao et al. (2024) in China, which revealed that ASA grade > II was an independent risk factor for anastomotic leak. This difference is explained by the fact that, the China study was a multi center study with a larger sample size compared to our study.(Gao et al., 2024).

In our study, emergency surgery was not a statistically significant risk factor for anastomotic leak. This is contrary to the study in Ethiopia by Zemenfes and Tamirat (2019), who found a significantly increased risk of leak following emergency surgery compared to elective surgery. This is likely due to the differences in the sample sizes of these studies. Zemenfes and Tamirat (2019) study, had a larger sample size (n=157) compared to our study with a smaller sample size in our study (n=85).

Low serum albumin < 3.5 g/dl wasn’t significant risk factor for anastomotic leak in our study. Other studies, however, contradict this study’s findings. A study in Egypt by Sieda et al. (2019), showed hypo-albuminemia < 3g/dl as a significant risk factor for anastomotic leak. In addition, Haxhirexha et al. (2019), showed that serum albumin level under 2.5 gm/dL was a significant risk factor for anastomotic leak after colorectal surgery. The reason for this variation is because the other 2 studies had lower cut off points ( < 2.5 g/dl and < 3 g/dl) for low serum albumin respectively, (Haxhirexha et al., 2019; Sieda et al., 2019); compared to our study which had a higher cut off point for low serum albumin ( < 3.5 g/dl).

### The effect of Hb on the occurrence of anastomotic leak

Haemoglobin level of less than 10g/dl was significantly associated with anastomotic leak. (RR, 8.15; 95% CI 1.16-57.48; p= 0.035). These findings are consistent with other studies in the literature. A prospective study in Indonesia on predictive factors for anastomotic leakage following hemicolectomy for colon cancer by Danardono et al. (2024), reported serum haemoglobin level ≤ 10.3 g/dl; a significant risk factor for anastomotic leak (OR = 3.4; p = 0.007). The Indonesian study had the same sample size as our study, (n=85).

Further more, another related study in Mulago by Nathani (2022), reported low hemoglobin <10g/dl; a significant risk factor for colorectal anastomotic leak. (OR = 6.25, p = 0.001).The reason for the similar findings between Nathani (2022) study and our study is likely due to similar study setting. Low hemoglobin causes reduced perfusion and oxygenation to the anastomotic ends which predisposes to poor healing and eventually an anastomotic leak.(Makanyengo et al., 2020).

However, raising Hemoglobin levels intraoperatively through blood transfusion is associated with increased anastomotic leak rates. A nation wide study in Denmark on anastomotic leakage after colonic cancer surgery by P□M Krarup et al. (2012), reported an association between intraoperative blood transfusion and anastomotic leak.(OR = 10.27; *p* < 0.001). Therefore, it may be beneficial to delay bowel anastomosis in elective cases, and for the emergency cases, placing end stomas may be required to reduce on the rate of anastomotic leak.

### Strengths of the study

The prospective cohort study allowed us to interact with participants and establish the factors associated with anastomotic leak.

The study cohort was homogenous and consisted of patients who underwent gastrointestinal anastomosis.

Confounding in this study was controlled by adjustment using multivariate logistic regression analysis.

### Limitations of the study

This study was done with a small sample size; hence, affected the reliability of our results, as shown by the wide confidence intervals for the relative risk of anastomotic leak.

## Conclusion

The incidence of anastomotic leak following gastrointestinal surgery was low (8.2%). Low Hemoglobin level, Hb < 10 g/dl was the only statistically significant predictor for gastrointestinal anastomotic leak. Other factors such as low serum albumin and cadre of surgeon (SHO) were associated with anastomotic leak but were not independent predictors of leaks. These findings support the importance of thorough preoperative patient assessment prior to gastrointestinal surgery.

## Recommendations

The surgical team should avoid primary bowel anastomoses in emergency gastrointestinal patients who present with low hemoglobin levels. These patients are prone to developing anastomotic leaks.

For elective cases, requiring secondary bowel anastomosis (Stoma reversals), the patient’s surgery should be deferred until the hemoglobin level has been raised to at least 10g/dl.

## Data Availability

All data produced in the present work are contained in the manuscript

## Acknowledgement

Dr. Paul Okeny, Dr. Ronald Mbiine and Dr. Wilberforce Musoga Kabweru for their support and guidance during this research. Special thanks to my research assistants, Biostatistician (Dr. Brian Kasagga) and Dr. Flavius Ebaisem Egbe for their considerable efforts in completing this work. I sincerely thank all the patients and their caregivers who unconditionally agreed to participate in this study.

## Notes

### Competing Interest Statement

The authors have declared no competing interest.

### Funding Statement

This study did not receive any funding

### Author Declarations

Makerere University School of Medicine Ethics committee gave approval for this work. Mak-SOMREC-2024-899

### Summary of Updates

The abstract and some sections within the methods were adjusted to include the exact dates of the participant recruitment and the form of informed consent obtained from the participants

